# Severe biventricular cardiomyopathy in both current and former long-term users of anabolic-androgenic steroids

**DOI:** 10.1101/2023.09.06.23295123

**Authors:** Rang Abdullah, Astrid Bjørnebekk, Lisa E. Hauger, Ingunn R. Hullstein, Thor Edvardsen, Kristina H. Haugaa, Vibeke M. Almaas

**Author notes:** **Address for correspondence:** Vibeke Marie Almaas, MD, PhD, Department of Cardiology, Oslo University Hospital, Rikshospitalet, Sognsvannsveien 20, 0372 Oslo, Norway / PO Box 4950 Nydalen, 0424 Oslo, Norway. **Institution where the work was performed:** Department of Cardiology, Oslo University Hospital, Rikshospitalet.

## Abstract

**Aims:** Explore the cardiovascular effects of long-term anabolic-androgenic steroid (AAS)-use in both current and former weightlifting AAS-users, and estimate the occurrence of severe reduced myocardial function and the impact of duration and amount of AAS.

**Methods:** In this cross-sectional study 101 weightlifting AAS-users with at least one year cumulative AAS-use (mean 11±7 accumulated years of AAS-use) were compared to 71 non-using weightlifting controls (WLC) using clinical data and echocardiography.

**Results:** Sixty-nine were current, 30 former (> 1 year since quitted), and 2 AAS-users were not available for this classification. AAS-users had higher left ventricular mass index (LVMI) (106±26 versus 80±15 g/m^2^, P<0.001), worse LV ejection fraction (LVEF) (49±7 versus 59±5%, P<0.001) and right ventricular global longitudinal strain (RVGLS) (−17.3±3.5 versus −22.8±2.0%, P<0.001), and higher systolic blood pressure (SBP) (141±17 vs. 133±11 mmHg, p<0.001) compared with WLC. In current users accumulated duration of AAS-use was 12±7 years, and in former 9±6 years (quitted 6±6 years earlier). Compared to WLC, LVMI and LVEF were pathological in current and former users (p<0.05) with equal distribution of severely reduced myocardial function (LVEF ≤40%) (11% vs. 10%, NS). In current users estimated life time AAS-dose correlated with reduced LVEF and LVGLS, p<0.05, but not with LVMI, p=0.12. Regression analyses of the total population showed that the strongest determinant of reduced LVEF were not coexisting strength training or hypertension, but history of AAS-use (β −0.53, P<0.001).

**Conclusions:** Long-term AAS-users showed severely biventricular cardiomyopathy. The reduced systolic function was also found upon discountied use.

## INTRODUCTION

Anabolic-androgenic steroids (AAS) constitute a wide range of synthetic derivatives of the male sex hormone testosterone that are primarily used in an illicit manner to gain muscle mass for cosmetic or ergogenic purposes.(1, 2) A meta-study in 2014 estimated the global lifetime prevalence of AAS-use at 6.4% among men, and 1.6% among women, suggesting that nonmedical AAS-use is widespread.(3)

Several case-reports, and small-scale studies have described an association between high-dose AAS-use and myocardial hypertrophy and hypertension.(4) In addition, post-mortem studies have examined young male bodybuilders who suffered sudden cardiac death, and found an association between AAS-use and myocardial hypertrophy and fibrosis.(4–7) It was not until the 2000s, that larger cross-sectional studies were published on the subject. Baggish et al. and later Fyksen et al. demonstrated an association between long-term AAS-use and myocardial hypertrophy, moderately decreased left ventricular (LV) systolic function, and coronary atherosclerosis.(8, 9) Furthermore, Rasmussen et al., compared current and former AAS-users and found moderately decreased LV systolic function in both groups, suggesting that the toxic cardiovascular effects are not fully reversible.(10) The underlying mechanisms of how supraphysiological doses of AAS induce adverse cardiovascular effects are not fully understood. Existing studies often include small numbers of participants, with varying lifetime duration of AAS-use and definitions of how to define AAS-use, including current and former use. Additionally, only few studies have properly adjusted for the use of psychostimulants, which are prevalent among users of AAS,(11) and associated with acute and long-term cardiovascular effects,(12–14). These factors may contribute to conflicting results, and as a result, an ongoing debate of what contributes to the adverse cardiovascular effects found among AAS-users.

The relationship between blood pressure and AAS-use has been debated. In the 2018 ESC/ESH clinical practice guidelines for the management of arterial hypertension, AAS abuse is listed as one of the causes for resistant hypertension.(9, 15–17) In a 2022 position paper on the cardiovascular effects of doping substances, it is still unclear whether pathological myocardial hypertrophy is attributed to arterial hypertension or to the direct effect of AAS on the myocardium.(18)

In this large scale cross-sectional study we explored the long-term effects of supraphysiological AAS-doses on myocardial hypertrophy and myocardial function, and if a substantial proportion of the users develop severely reduced myocardial function, by comparing current and former AAS-using weightlifters with a non-using weightlifting control (WLC) cohort, employing echocardiographic imaging techniques. We hypothesized that AAS-use is the strongest independent determinant of both increased myocardial hypertrophy and decreased function, and that increased duration and lifetime doses of AAS is associated with worsened effects.

## METHODS

### Study design and population

We included men involved in heavy strength training, who volunteered from the community to participate in this cross-sectional study from May 2017 to October 2019. We recruited participants through social media, targeted online fora, and posters and flyers distributed in select gyms in Oslo, Norway. The participants were compensated with a 500 NOK (approximately $60) gift card for their participation. The study is part of a longitudinal study investigating the impact of high-dose AAS-use on the brain, carried out at the Anabolic Androgenic Steroid Research Group at Oslo University Hospital, Norway (https://www.ous-research.no/anabolic-steroids/). The participants were recruited from this project, and the sample is partly overlapping with the one described in previous brain health publications.(16, 19, 20)

A self-reported one repetition maximum (1RM) bench press of at least 100 kg (approximately 220 lbs) was required to enter the study. The participants were either current or former (defined as at least one year since cessation of AAS-use) AAS-users reporting at least one year of cumulative AAS-use, or men who had never been exposed to AAS or equivalent doping substances (weightlifting controls, WLC). In addition, WLC who were treated medically for hypertension were excluded. The study was conducted in accordance with the declaration of Helsinki and received ethical approval from the Regional Committee for Medical and Health Research Ethics in South-Eastern Norway (REK) (2013/601). Participants received an informational brochure with a complete description of the study prior to participation, and written informed consent was obtained from all participants.

### Screening instruments, clinical interview and examination

Self-report questionnaire, and a semi-structured clinical interview, performed by two investigators (AB, LH) was used to assess relevant background and health information, including hours spent per week on strength and endurance training, weekly intake of alcohol (in units), history of smoking and use of illicit drugs. In particular, current or former use of the psychostimulants cocaine and amphetamines were mapped with questions related to whether they had ever used the substances (yes/no) and a follow-up quantification question, which was later combined to a 0-4 scale, with the options; never used, less than once a month, monthly, weekly and daily/almost daily. In addition, AAS-users were interviewed about the history and nature of AAS-use, including AAS substances used, and whether and when they had ceased using AAS. Estimated lifetime AAS dose was calculated as the lifetime average weekly dose reported and lifetime weeks of AAS exposure, in line with previous studies.(20, 21) We obtained blood pressure measurements using an automatic sphygmanometer, with a standard bladder cuff for most patients, and larger cuffs for bigger arm circumferences.

### Doping analysis

Urine samples were collected and analysed for external use of androgens using gas and liquid chromatography coupled to mass spectrometry at the WADA accredited Norwegian Doping Laboratory at Oslo University Hospital.(22) The criteria used to determine external androgen use were: 1) urine samples positive for synthetic testosterone compounds 2) a testosterone to epitestosterone ratio (T/E) >15 equivalent to previous work.(22–25) To minimize the problem of some participants not disclosing their use of AAS, WLC with urine findings suggestive of AAS or image and performance enhancing drugs banned by the World Anti-Doping Agency (WADA)(26) were excluded from further evaluation.

### Transthoracic echocardiography

Echocardiography was performed using Vivid E95 (GE Vingmed Ultrasound, Horten, Norway). All echocardiographic measurements were obtained by one investigator (VMA) and analysed offline, blinded to AAS status, by another investigator (RA) using the software EchoPAC v203 (GE, Horten, Norway). Two-dimensional (2D) and Doppler echocardiographic measurements were performed according to current standards.(27) LV ejection fraction (LVEF) was calculated by modified Simpson’s biplane method (LVEF Simpson). LV systolic function was further assessed by global longitudinal strain (LVGLS) using speckle-tracking in the three apical views and defined as the average of peak longitudinal strains from a 16 LV segments model. If two or more segments in any given view were deemed inappropriate for strain assessments, LVGLS was excluded. Diastolic function was assessed by transmitral pulsed Doppler in the apical four-chamber view by measuring peak mitral inflow velocities of early and late atrial filling (E-wave and A-wave). At the mitral annulus, peak early velocity (e’) was measured at both septal and lateral locations. Average e’ was then calculated from these two measurements. We calculated left atrial end-systolic volume using the area length method in apical four- and two-chamber views. Maximal wall thickness (MWT) was assessed from parasternal short-axis view from all LV segments from base to the apex of the LV. LV mass was estimated from parasternal views using the formula provided by Devereaux et al.(28) Right ventricular (RV) linear dimensions were estimated from a RV-focused apical four-chamber view. The proximal RV outflow tract (RVOT) diameter was measured in the parasternal long-axis. RV systolic function was evaluated using two parameters: fractional area change (FAC) and right ventricular global longitudinal strain (RVGLS). FAC was obtained in a RV-focused apical four-chamber view and calculated from the difference in end-systolic and end-diastolic area divided by the area in diastole and expressed as a percentage. RVGLS was obtained using speckle-tracking.

### Statistical analyses

Continuous data are presented as mean±standard deviation (SD) or median (25th-75th percentile). Student’s t-test were used to compare means between two groups, and ANOVA and Bonferroni post hoc test when comparing three groups. Proportions were compared by the Pearson’s χ^2^ or Fischer’s Exact test. Linear regression models were used to assess the strength of the relationship between the continuous variables LV mass indexed to BSA (LVMI), LVEF, LVGLS and RVGLS and predictor variables known to impact cardiac function or structure such as age, use of AAS, weekly hours of resistance and endurance training and systolic blood pressure (SBP). Results were presented as standardized beta coefficient (ß) (95% CI), and significance level (P value). Significant predictors (defined as p<0.10) from univariate models with no adjustment for covariates were included in a multivariate model, with the exception of age, which was forced in.

#### Sensitivity analyses

To further evaluate possible differences between WLC, current and former AAS-users, similar linear regressions models were run with LVMI, LVEF, LVGLS, and RVGLS as dependent variables, group as fixed factor, age and the significant predictors (bench press 1RM, SBP and LVMI for functional measures) included as covariates in the model. Bonferroni post hoc tests were used to test for differences between the three groups. Also, since use of stimulants are known to cause adverse cardiovascular effects,(12, 13) additional sensitivity analyses were computed including ordinal measures of amphetamine and cocaine use as additional covariates in these models. These were run as separate sensitivity analyses as more data were missing on these variables, to preserve power in the main model. Lastly, to test for possible effects of degree of AAS exposure, the regression model was rerun comparing WLC and subgroups of current AAS-users that differ in their duration of AAS-use (<5, 5-10, and >10 years of use). In addition, the relations among between LVMI, LVEF, and LVGLS and estimated lifetime AAS-dose were investigated with Pearson’s or Spearman’s correlations upon violation of normality and/or linearity among current AAS-users. All statistical analyses were performed using SPSS version 25.0 (SPSS, Chicago, Illinois, USA).

## RESULTS

### Clinical characteristics

#### AAS-users versus WLC

We included 172 participants, 101 were AAS-users and 71 WLC. The accumulated duration of AAS-use was 11±7 years. AAS-users had higher weight (P=0.003), BMI (P=0.002), and BSA (P=0.02) compared with WLC (Table 1). SBP was higher in AAS-users compared with WLC (P<0.001), while diastolic blood pressure was similar (P=0.12). None of the WLC were medically treated for hypertension, nor did they test positive for AAS or had a T/E ratio above threshold. The frequencies of the specific AAS compounds found in the urine samples are summarized in Supplemental Figure 1 along with a summary of the most commonly used compounds based on self-reports. In addition to AAS, 10% of users reported use of growth hormones and/or clenbuterol in current or last cycle, and 18% reported use of post cycle supplement drugs such as selective estrogen receptor modulators, aromatase inhibitors and/or human chorionic gonadotropin. Post cycle therapy is commonly used following an AAS cycle to prevent further conversion of excessive testosterone into oestrogens via the aromatase enzyme, to restore the hypothalamus-pituitary-gonadal axis and to recover endogenous testosterone production.

**Table 1.**
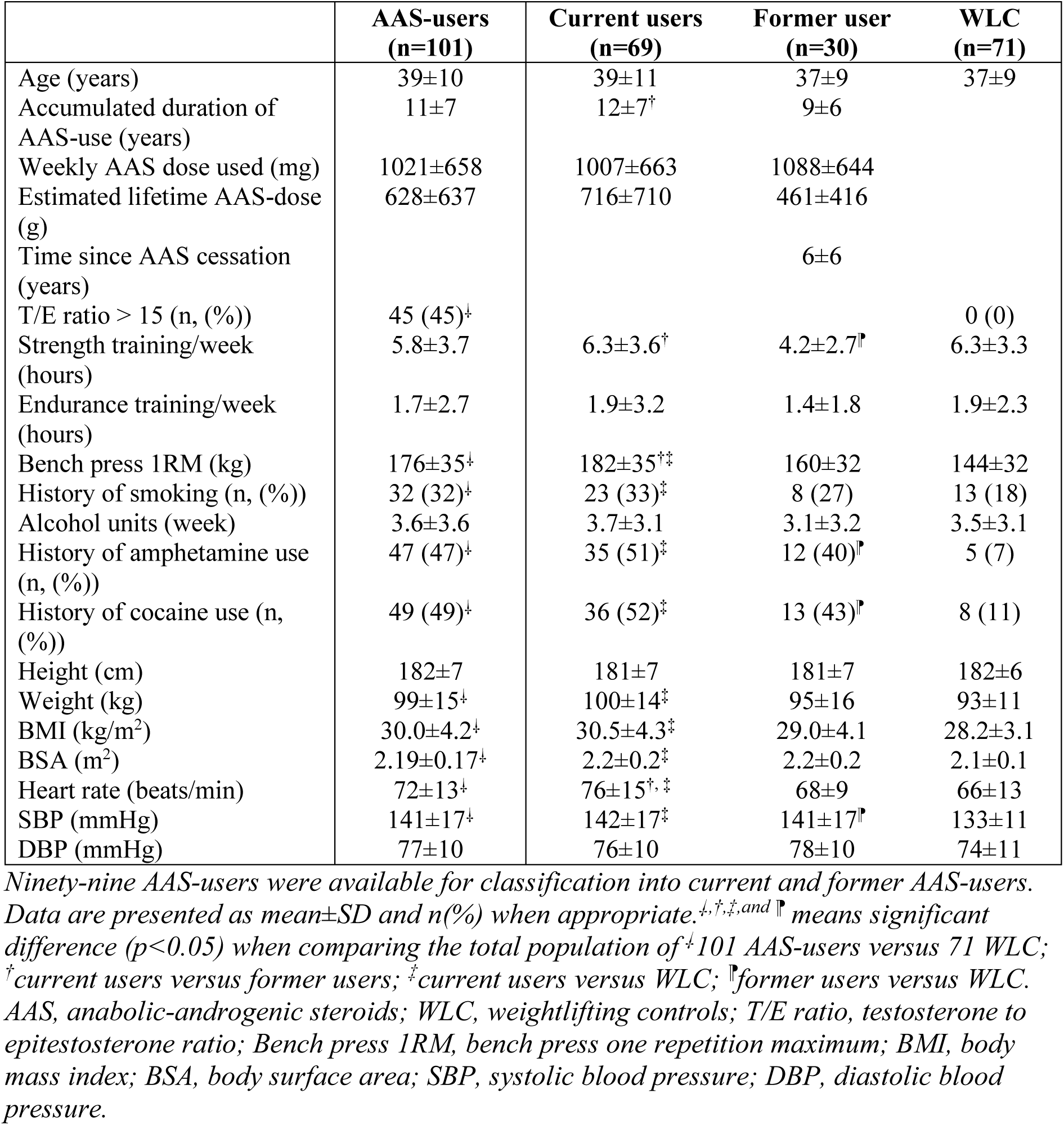
Comparisons of clinical characteristics in the total population of 101 AAS-users and in current or former AAS-users, with 71 non-using WLC.

|  | AAS-users<br>(n=101) | Current users<br>(n=69) | Former user<br>(n=30) | WLC<br>(n=71) |
| --- | --- | --- | --- | --- |
| Age (years) | 39±10 | 39±11 | 37±9 | 37±9 |
| Accumulated duration of<br>AAS-use (years) | 11±7 | 12±7<sup>†</sup> | 9±6 |  |
| Weekly AAS dose used (mg) | 1021±658 | 1007±663 | 1088±644 |  |
| Estimated lifetime AAS-dose<br>(g) | 628±637 | 716±710 | 461±416 |  |
| Time since AAS cessation<br>(years) |  |  | 6±6 |  |
| T/E ratio > 15 (n, (%)) | 45 (45)<sup>‡</sup> |  |  | 0 (0) |
| Strength training/week<br>(hours) | 5.8±3.7 | 6.3±3.6<sup>†</sup> | 4.2±2.7<sup>¶</sup> | 6.3±3.3 |
| Endurance training/week<br>(hours) | 1.7±2.7 | 1.9±3.2 | 1.4±1.8 | 1.9±2.3 |
| Bench press 1RM (kg) | 176±35<sup>‡</sup> | 182±35<sup>†‡</sup> | 160±32 | 144±32 |
| History of smoking (n, (%)) | 32 (32)<sup>‡</sup> | 23 (33)<sup>‡</sup> | 8 (27) | 13 (18) |
| Alcohol units (week) | 3.6±3.6 | 3.7±3.1 | 3.1±3.2 | 3.5±3.1 |
| History of amphetamine use<br>(n, (%)) | 47 (47)<sup>‡</sup> | 35 (51)<sup>‡</sup> | 12 (40)<sup>¶</sup> | 5 (7) |
| History of cocaine use (n,<br>(%)) | 49 (49)<sup>‡</sup> | 36 (52)<sup>‡</sup> | 13 (43)<sup>¶</sup> | 8 (11) |
| Height (cm) | 182±7 | 181±7 | 181±7 | 182±6 |
| Weight (kg) | 99±15<sup>‡</sup> | 100±14<sup>‡</sup> | 95±16 | 93±11 |
| BMI (kg/m<sup>2</sup>) | 30.0±4.2<sup>‡</sup> | 30.5±4.3<sup>‡</sup> | 29.0±4.1 | 28.2±3.1 |
| BSA (m<sup>2</sup>) | 2.19±0.17<sup>‡</sup> | 2.2±0.2<sup>‡</sup> | 2.2±0.2 | 2.1±0.1 |
| Heart rate (beats/min) | 72±13<sup>‡</sup> | 76±15<sup>†, ‡</sup> | 68±9 | 66±13 |
| SBP (mmHg) | 141±17<sup>‡</sup> | 142±17<sup>‡</sup> | 141±17<sup>¶</sup> | 133±11 |
| DBP (mmHg) | 77±10 | 76±10 | 78±10 | 74±11 |

Ninety-nine AAS-users were available for classification into current and former AAS-users. Data are presented as mean±SD and n(%) when appropriate. <sup>‡</sup>, <sup>†</sup>, <sup>‡</sup>, and <sup>¶</sup> means significant difference ( $p<0.05$ ) when comparing the total population of <sup>‡</sup>101 AAS-users versus 71 WLC; <sup>†</sup>current users versus former users; <sup>‡</sup>current users versus WLC; <sup>¶</sup>former users versus WLC. AAS, anabolic-androgenic steroids; WLC, weightlifting controls; T/E ratio, testosterone to epitestosterone ratio; Bench press 1RM, bench press one repetition maximum; BMI, body mass index; BSA, body surface area; SBP, systolic blood pressure; DBP, diastolic blood pressure.

#### Current, former and no AAS-use

Among 101 AAS-users, 69 were current users, 30 were former users, and 2 users were unclassifiable with regard to current or former status, and therefore not included in further subgroup analyses (Table 1). Mean time since AAS cessation among former users was 6±6 years (median 3.75 (1–25)). Former AAS-users reported approximately 3 years shorter accumulated duration of AAS-use (9±6 vs. 12±6 years, P=0.05), but estimated lifetime AAS-dose did not significantly differ between former and current users (461±416 vs. 716±710 g, P=0.07). SBP was also at the same high level in former and current users, significantly different from WLC. Doping analyses tests indicative of AAS-use were seen in 81% (n = 53) of current users and in 17% (n = 3) of former users. The positive tests among former users involved elevated ratio between testosterone to epitestosterone (T/E ratio) and were consistent with reported medical use of testosterone replacement therapy.

### Echocardiography

#### AAS-users versus WLC

Measurements of LV structure were consistent with AAS-users displaying concentric hypertrophy, with markedly higher LVMI, MWT and relative wall thickness (all P<0.001) compared with WLC (Table 2, Figure 1 panel A). AAS-users had reduced LV function, by LVEF and LVGLS compared with WLC (Table 2). Of note, 36% of AAS-users had LVEF between 41-49%, and 11% had LVEF below or equal to 40%. RV systolic function measured by RVGLS and FAC was impaired among AAS-users compared with WLC. Assessment of LV diastolic function according to the ASE/EACVI guidelines (29) showed that only three AAS-users (3%) were found to have grad II diastolic dysfunction, and no users in grad I or III. (Table 2). Like all the WLC, 97% (98/101) of the AAS-user did not have diastolic dysfunction according to ASE/EACVI guidelines.

**Tables 2.**
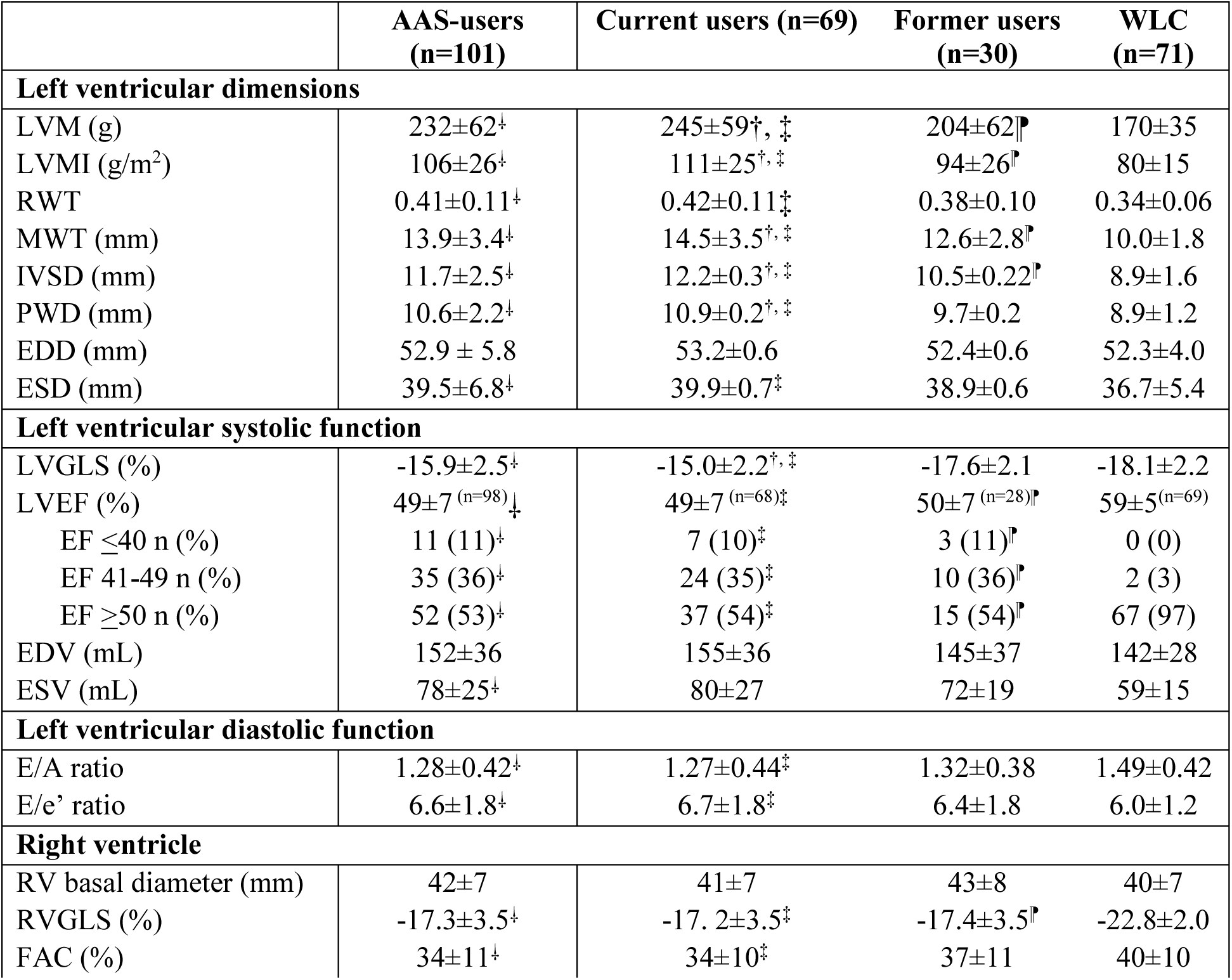
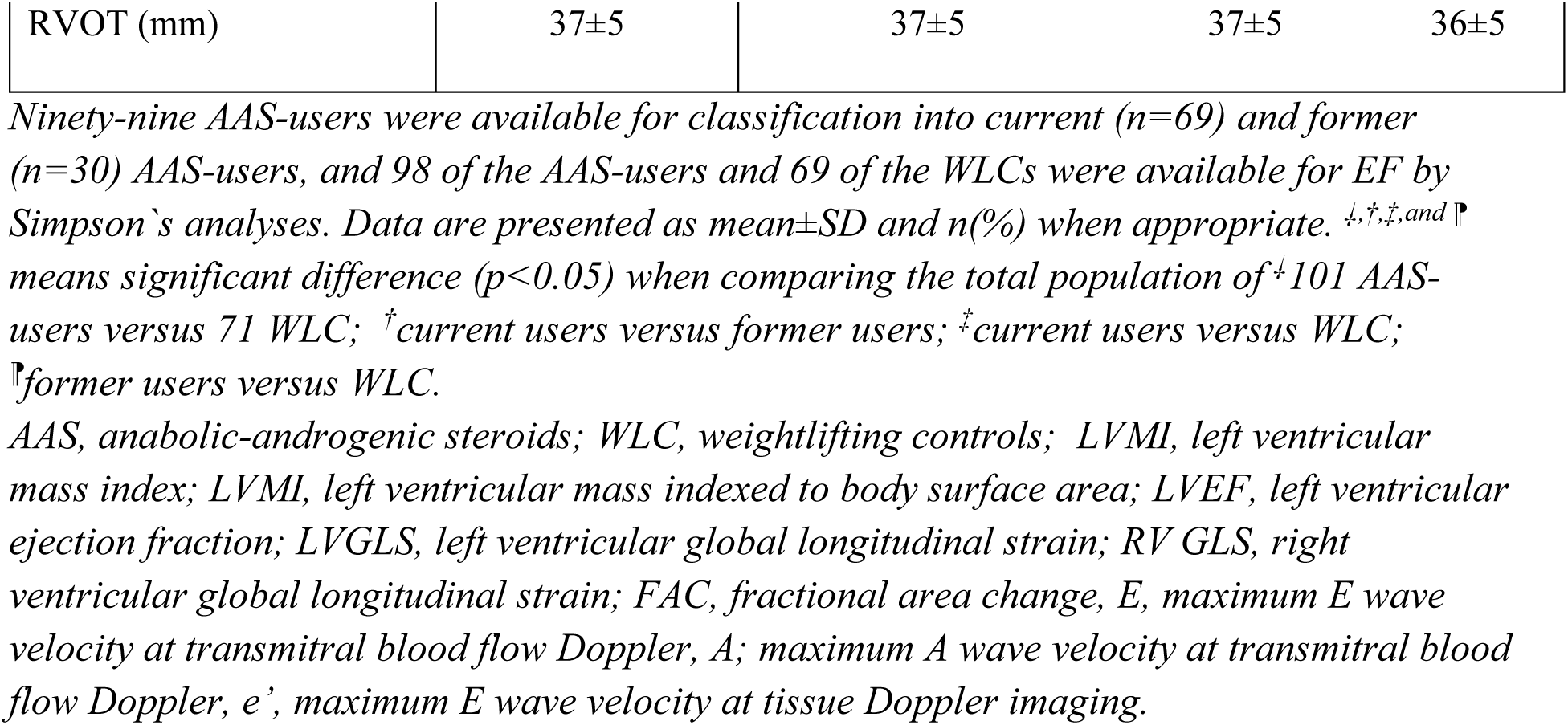
Comparisons of echocardiographic parameters in the total population of 101 AAS-users and in current or former AAS-users, with 71 non-using WLC.

**Figure 1.**
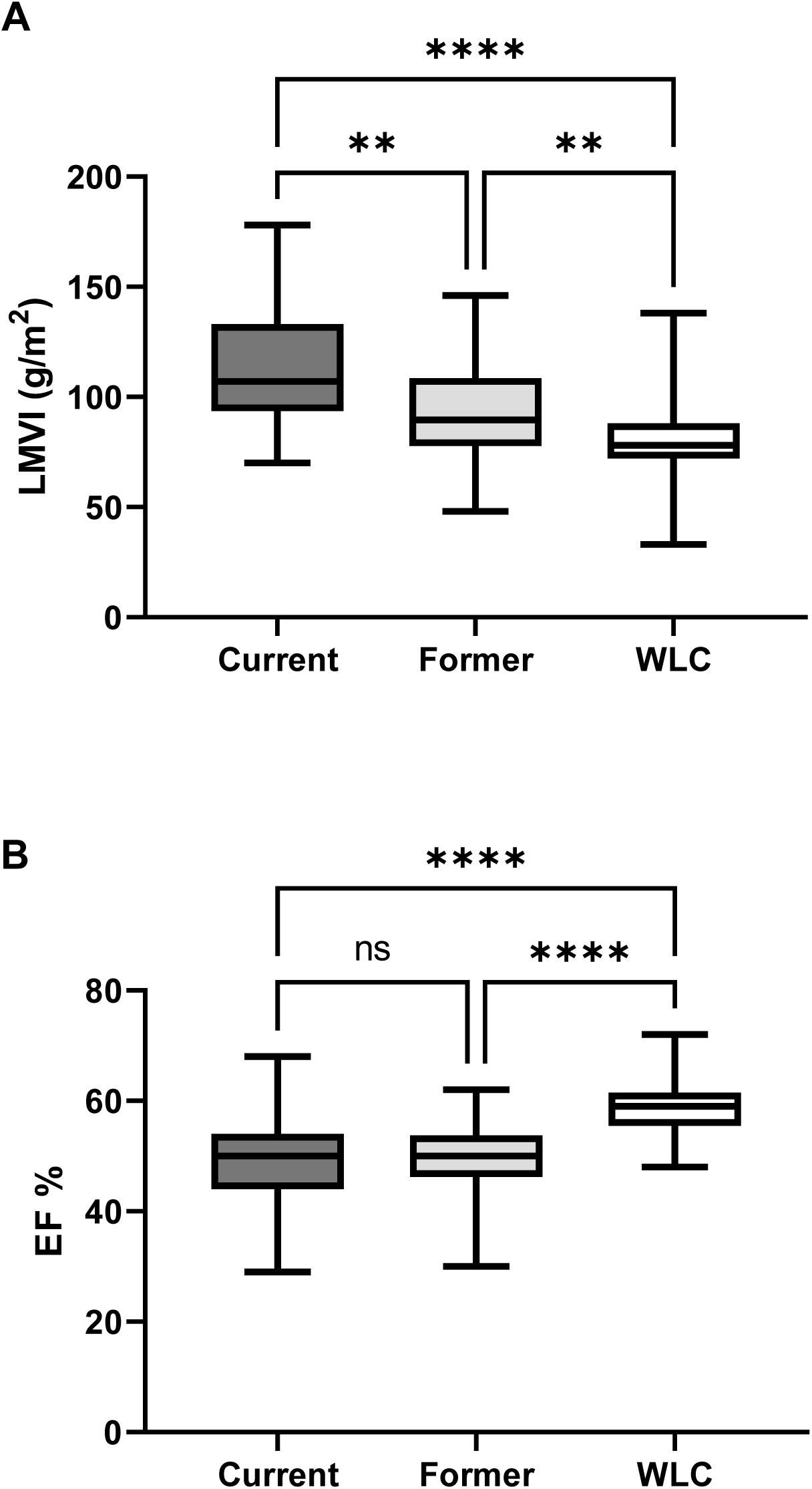
Box-plot showing myocardial hypertrophy and myocardial function in current and former AAS-users and WLC. Panel A) shows boxplots of LVMI in current AAS-users, former AAS-users and WLC. On this variable, LVMI was 111±26 g/m^2^ among current users, 98±26 g/m^2^ among former users, and 80±15 g/m^2^ among WLC. Panel B) shows EF in the same 3 groups. On this variable, EF was 49±7% among current users, 50±6% among former users, and 59±5% among WLC. Ns means not significant, **p<0.01 and ****p<0.001. AAS, anabolic androgenic steroid; WLC, weight lifting controls; LVMI, left ventricular mass index; EF, ejection fraction.

#### Current, former and no AAS-use

Current AAS-use was associated with greater pathology, whereas former users were somewhere in between current users and WLC (Table 2, Figure 1, Supplemental Table 1). The proportion of current and former users by LVEF category (≤40%, 41-49 % and ≥50%) did not significantly differ (Table 2).

#### Severely reduced left ventricle ejection fraction among AAS-users

The distribution between severely reduced LVEF (LVEF≤40%), moderat reduced LVEF (LVEF 41-49%) and subnormal or normal LVEF (LVEF ≥ 50%) are showed in Table 2 and illustrated in Figure 2. The 11% AAS-users with severely reduced LVEF (LVEF ≤ 40%) had higher weight and BMI, and higher weekly AAS dose compared to the AAS-users with moderate reduced or subnormal/normal LVEF, but estimated lifetime AAS dose, strength training load and bench press 1RM did not achieve significance (Table 3).

**Figure 2.**
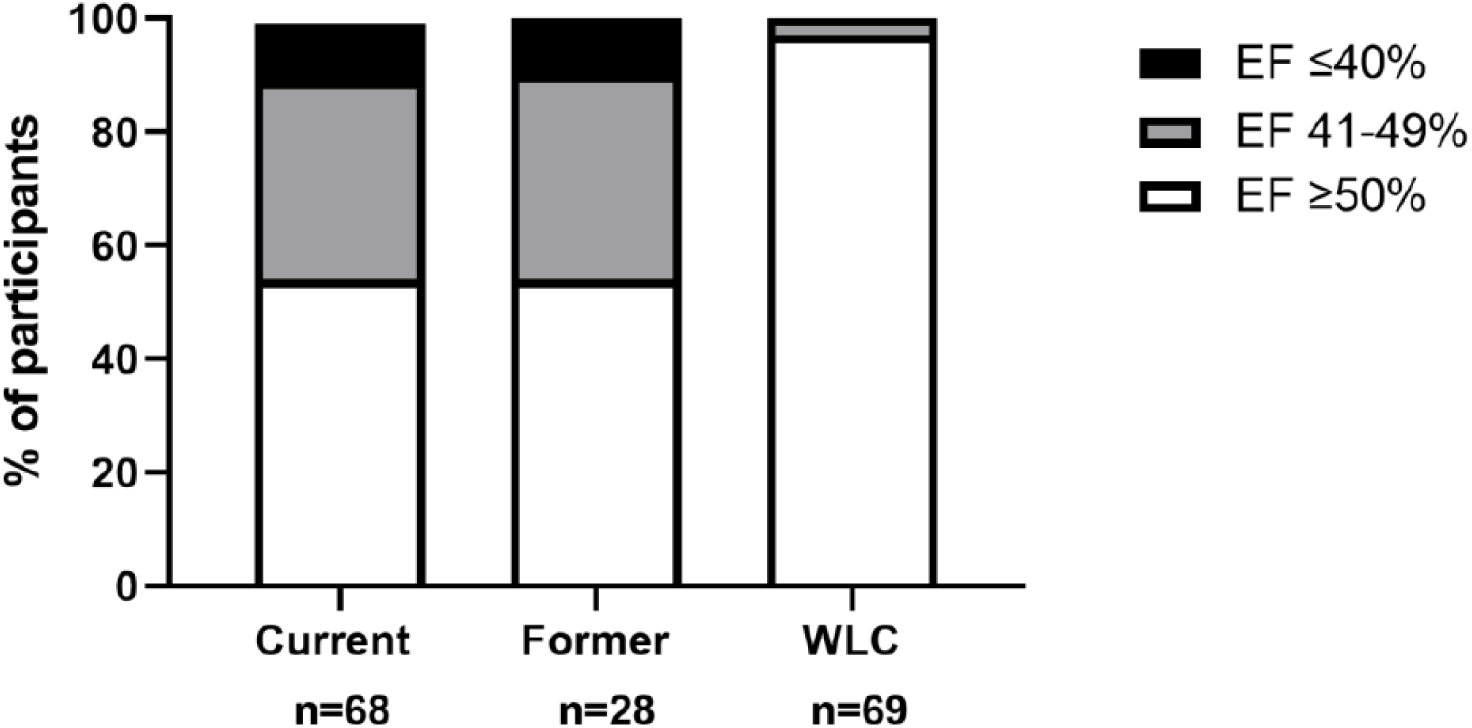
Proportions of severely and moderately reduced ejection fraction and subnormal or normal ejection fraction in current and former AAS-users and WLC. Severely (EF<41%) and moderately (EF 41-49%) reduced ejection fraction and subnormal or normal (EF>49%) ejection fraction in current and former AAS-users and non-using WLC. Of 101 AAS-users, 68 current and 28 former AAS-users were available for EF Simpsons-analyses and of 71 weightlifting controls 69 were available for EF Simpsons-analyses. AAS, anabolic androgenic steroid; EF, ejection fraction; WLC, weightlifting controls.

**Table 3.**
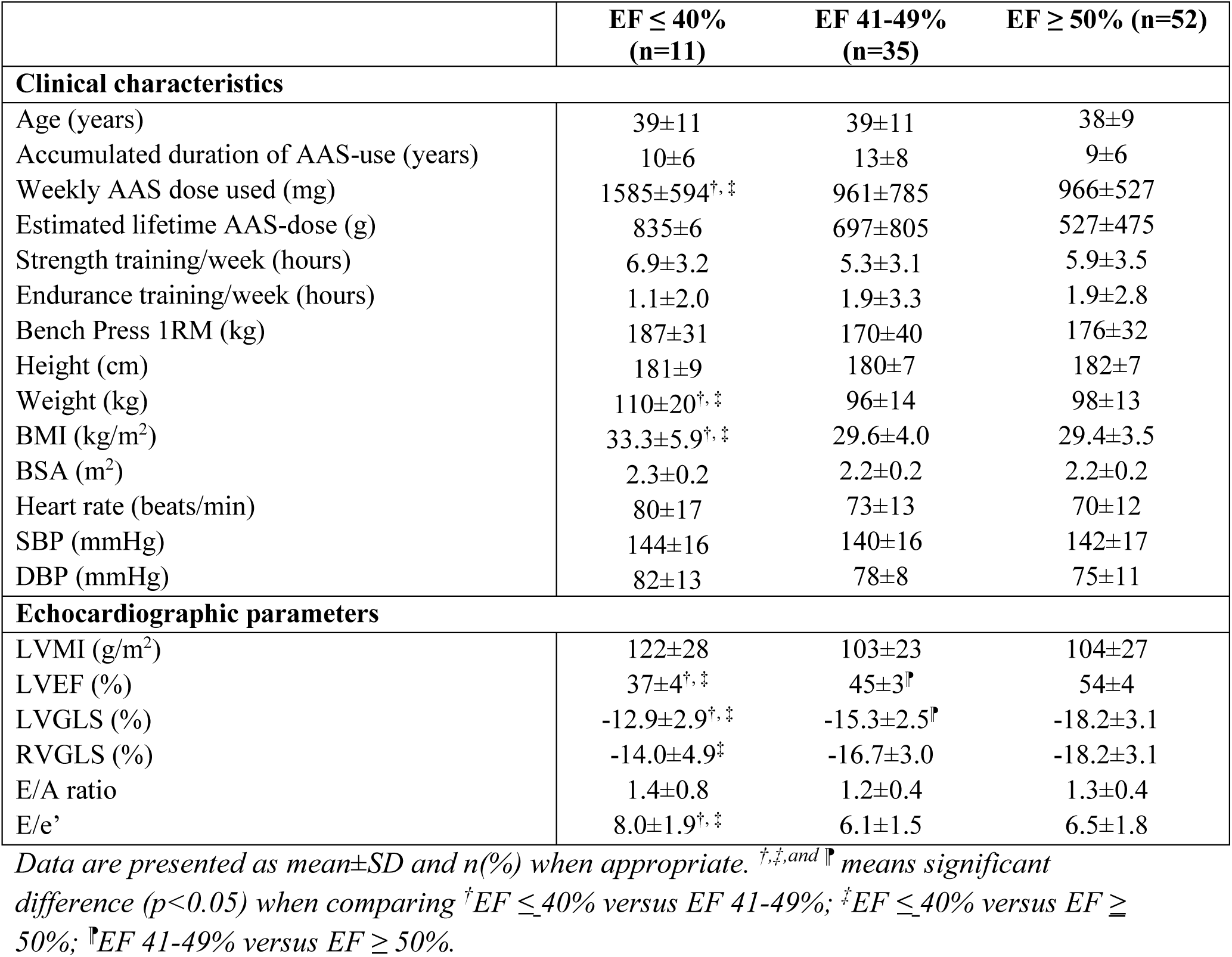
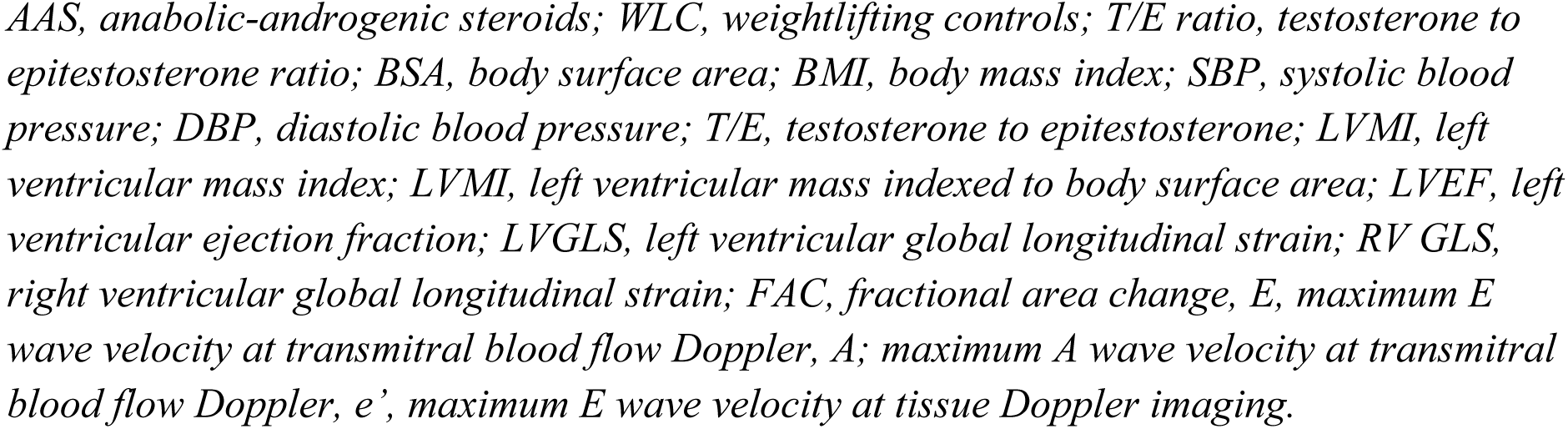
Clinical characteristics and a set of key echocardiographic parameters in AAS-users with EF ≤ 40%, EF 41-49%, and EF ≥ 50%.

#### Linear regression analysis for determinants of biventricular cardiomyopathy in the total population

In the multivariate regression model, history of AAS-use was the strongest independent determinant of higher LVMI, while strength training, SBP and bench press 1RM contributed to a lesser degree to the model (Supplemental Table 2). History of AAS-use was the only independent determinant of decreased LVEF and of reduced RV function (Supplemental Table 3 and 5). LVMI was identified as the strongest independent determinant of reduced LV function, closely followed by history of AAS-use (Supplemental Table 4). Sensitivity analyses revealed that the main effect of group remained significant for the two main measures of LV function, LVEF and LVGLS, when amphetamine and cocaine were included as additional covariates in these models (Supplemental Table 6 and 7).

#### Impact of duration of use on key echocardiographic parameters

Sensitivity analyses among current AAS-users divided into subgroups based on duration of AAS-use, showed main effects of group on all echocardiographic parameters (Supplemental Table 8). The post hoc comparisons indicated that all significant differences applied to differences between the WLC and subgroups of AAS-users, whereas no significant differences were found between subgroups of users varying in different length of AAS-use. Briefly, for LVMI and RVGLS, all AAS subgroups were significantly different compared to the WLC, with higher mass or reduced GLS, respectively. For LVEF and LVGLS significant differences were also found, although not after shorter exposure of AAS (<5 years) (Supplemental Table 8). Lastly, Spearman’s “Rho” correlations, including only current AAS-users (n=69), showed significant relations between estimated lifetime AAS-dose and LVEF (r=-30.7, p=0.01, n=67), and LVGLS (r=30.7, p=0.01, n=50), but no relation to LVMI (r=0.19, p=0.12, n=68) (Figure 3).

**Figure 3.**
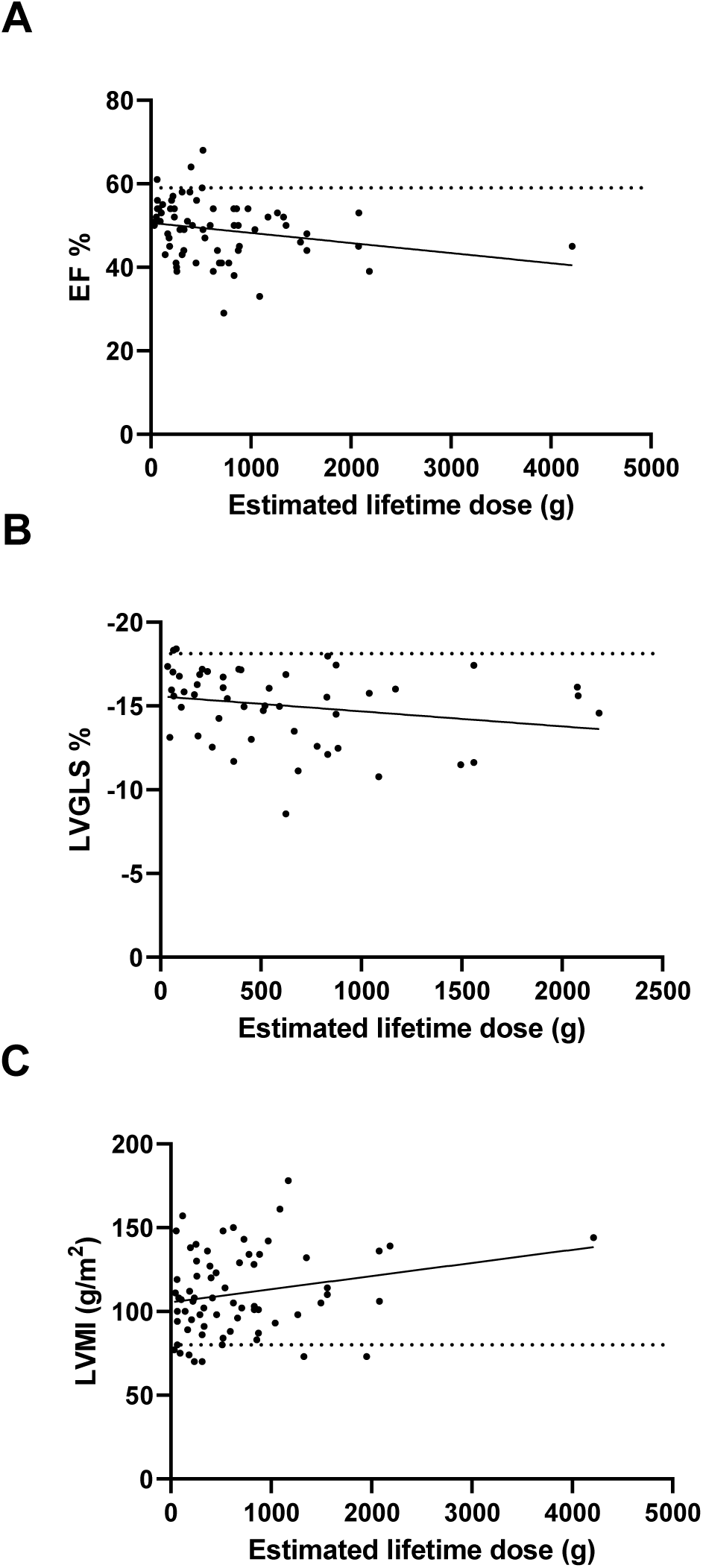
Scatterplot showing correlations between estimated lifetime AAS-dose and myocardial function and myocardial hypertrophy in 69 current AAS-users. Spearman’s “Rho” correlations in 69 current AAS-users showed significant relations between estimated lifetime AAS-dose and EF (r=-30.7, p=0.01, n=67) (A), and LVGLS (r=30.7, p=0.01, n=50) (B), but no relation to LVMI (r=0.19, p=0.12, n=68) (C). Dotted lines show mean value for EF (59±5%), LVGLS (−18.1±2.2%) and LVMI (80±15g/m^2^) in WLC. AAS, anabolic androgenic steroid; EF, ejection fraction; LVGLS, left ventricle global longitudinal strain; LVMI, left ventricle myocardial mass indexed for body surface area; WLC, weightlifting controls.

## DISCUSSION

This study represents the largest cardiovascular study to date, comparing AAS-users to WLC, enabling subgroup analyses to be performed on AAS-users with adverse pathology. There are several important aspects to consider. First, our study showed that long-term AAS-use was strongly associated with left ventricular hypertrophy (LVH), reduced LV and RV systolic function, and impaired diastolic function indicating that long-term AAS-use is associated with biventricular cardiomyopathy. Second, increased estimated lifetime AAS dose and duration of AAS-use was associated with increased myocardial pathology. Third, history of AAS-use was the strongest independent determinant of increased LVH and decreased LV systolic function, and outperformed psychostimulants, SBP and strength training. Fourth, former AAS-users had reduced LV and RV systolic function and sustained increased SBP, comparable to that of current AAS-users. Finally, 11 % of the AAS-users had severely reduced LVEF <40%.

### Cardiac structure and function

Using sensitive echocardiographic tools, we were able to demonstrate that long-term AAS-users when compared with WLC, exhibited cardiovascular changes compatible with a biventricular cardiomyopathy. AAS-users had thicker LV walls and a higher LVMI, reflecting the structural hypertrophic changes that are associated with AAS, consistent with findings in past studies.(8, 10) The rather moderate increase in LV wall thickness in our WLC, can be explained by the blood pressure response and cardiac output during weightlifting, as heavy-resistance training increases blood pressure up to 480/350 mmHg.(30) The moderate LVH is also found among other strength athlete cohorts, while LV systolic function generally remains unchanged.(31)

LV systolic function, as assessed by both LVEF and LVGLS, was significantly impaired in AAS-users compared to WLC, with substantial absolute differences, also consistent with past studies.(8, 10) The findings could not be explained by concurrent psychostimulant use, albeit we cannot rule out that co-current use would present an additional risk for acute cardiovascular events. Importantly, our study featuring a large population, provided a unique opportunity to examine previously unexplored subgroups of interest. Specifically, we observed a high occurrence of moderately and severely impaired LV systolic function (defined as LVEF 41-49% and ≤40% respectively) among both current and former AAS-users. A study combining two extensive community cohorts, found a five-year mortality rate of 66%, with 63% of deaths attributed to cardiovascular disease, following diagnosis of heart failure among patients with a reduced left ventricle ejection fraction (defined as LVEF <50%).(32) Our study adds important knowledge regarding the substantial risk of developing advanced heart failure upon long-term use of AAS, suggesting that AAS-users may be at high risk of cardiac death.

RV function was worse in AAS-users compared with WLC, as demonstrated by a lower FAC and RVGLS, and is consistant with the findings of D’Andrea et al,.(33) On RV linear dimensions and RVOT, both AAS-users and WLC had values either in the upper range or higher according to ASE/EACVI guidelines.(34) However, RV linear dimensions nor RVOT differed between AAS-users and WLC, which may reflect chronic maladaptive cardiac remodelling caused by weightlifting. RVOT is a frequent focus of ventricular tachycardia both in the general population and in athletes,(35) and is therefore of particular interest. In endurance athletes, lifetime exercise duration has been demonstrated to be associated with larger RVOT diameter.(36)

Our findings of myocardial hypertrophy in former users falls somewhere in between current users and WLC, suggesting a partial regression of hypertrophy following discontinuation. In contrast, reduced left and right systolic function measured with LVEF and RVGLS were at the same level in former and current AAS-users. Considering that the study sample comprises former AAS-users, that on average ceased several years ago, the persistently impaired biventricular myocardial function and increased systolic blood pressure is worth noting.

Our finding of reduced LV systolic function in former AAS-users, is in contrast with recent studies that have found such impairment selectively in current AAS-users. (8, 10, 37). In our study nearly half of the AAS-users, including both current and former users, had LVEF below 50%, and 11% LVEF below 40%, suggestive of moderate or severe LV systolic dysfunction in both current and former users. Our sample differs from previous studies in terms of the duration of AAS usage, showing an accumulated use of 12 years among current users and 9 years among former users. These durations notably exceed the corresponding durations observed in the aforementioned studies. Hence it is possible that the low LVEF of past users of AAS is a consequence of long-term use.

Among current AAS-users reporting <5 years, 5-10 years and >10 years of accumulative use, we found comparable levels of myocardial pathology in subsequent subgroup analyses. For LVMI and RVGLS, significant differences were observed between AAS-users with <5 years of use compared with WLC. While it is possible to infer that LVH and RV systolic dysfunctions may serve as early indications of cardiovascular toxicity associated with AAS usage, it is important to note that the limited sample size within the former subgroup hampers a definitive conclusion. Significant and more pronounced outcomes were observed with prolonged AAS usage among current users, indicating a progressive decline in myocardial function over time. These findings were supported by the observed correlation between decreased LVEF and LVGLS, and the increasing estimated lifetime AAS dosage.

Nevertheless, the findings point to long-term complications of use, suggesting persistent increased risk of adverse cardiovascular events, and needs to be further examined in large-scale longitudinal samples.

### Determinants of cardiac structure and function

The results of our univariate and multivariate linear regression analyses found that history of AAS-use was the strongest determinant of LVH measured by LVMI, and of LV and RV systolic function measured by LVEF and RVGLS respectively. History of AAS-use outperformed both SBP and strength training per week. It is not entirely clear whether the drug-mediated cardiovascular effects of AAS are secondary to that of hypertensive effects of AAS, or if AAS affects both blood pressure and cardiac parameters, although our linear regression analysis suggest that the latter may be true. Further longitudinal studies are necessary to evaluate susceptibility, progression, and independent predictors of LVH and decreased LV and RV systolic function.

### Clinical implications

This study showed that long-term use of AAS was associated with pathology affecting both ventricles, and thus compatible with a biventricular cardiomyopathy. The biventricular changes may be early manifestations of AAS-use. We showed that history of AAS-use outperformed hypertension and strength training, as the biggest contributor to cardiovascular pathology in users of AAS, in the largest cardiovascular study to explore determinants of pathology in AAS-users. We recruited AAS-users with a broad sociodemographic background from the general population, and not a subset of elite athletes. For that reason, clinicians who encounter long-term users of AAS should be aware that elevated blood pressure, LVH and impaired LV and RV systolic function, are not uncommon findings. Our study reiterates the critical need for longitudinal studies. It has been demonstrated that regression of LVH in hypertensive patients is associated with lower likelihood of cardiovascular morbidity and mortality.(41) A similar effect could occur in AAS patients upon cessation of substance use, but our study indicate that long term former AAS-use still show cardiovascular pathology several years after cessation of AAS-use.

### Limitations

This study has several limitations. First, this was a single centre, cross-sectional study, which does not allow claims of causality. Second, the exercise history and history and nature of AAS-use was self-reported and therefore subject to recall bias. The reported strength training per week did not take into account the lifetime training burden, yielding possible underestimates of athletic training on myocardial hypertrophy and function. Third, there are potential threats to the study’s generalizability. Although we recruited participants from the community, selection bias could still arise. Our study population consisted mainly of white Scandinavians, and therefore our results might not cover the full ethnic spectrum of AAS-users. Conversely, this yields in a high degree of racial homogeneity, meaning race will not interfere as a confounding factor in explaining cardiovascular differences between our two main cohorts of AAS-users and WLC. Finally, further evidence of an AAS-induced cardiomyopathy would require a longitudinal follow-up.

### Conclusions

Long term AAS-users exhibited myocardial hypertrophy, impaired LV and RV systolic function, and impaired diastolic function indicating a biventricular cardiomyopathy, with high occurrence of severely reduced ejection fraction. Importantly, substantial impairment of systolic function, as assessed by LVEF, and pathological increased systolic blood pressure were also seen in former users years after AAS discontinuation, which may indicate permanent changes in function. History of AAS-use was the strongest independent determinant of both LVH and reduced LV and RV systolic function. If not acted upon, high-dose AAS-use could prove to be a major public health concern with respect to adverse cardiovascular events.

## ACKNOWLEDGEMENTS

The authors are grateful for the valuable contributions made by all the study participants.

## Funding

This research was funded by the South-Eastern Norway Regional Health Authority (Grant Nos. 2016049, 2017025, 2018075 and 2020088 [to AB]), and by Norwegian Research council, #309762 Precision Health Center for optimized cardiac care (ProCardio).

## Conflict of interest

The authors report no biomedical financial interests or potential conflicts of interest.

## Data availability statement

The data underlying this article cannot be shared publicly due to privacy for the individuals that participated in the study. The data will be shared on reasonable request to the corresponding author.

## Notes

### Competing Interest Statement

The authors have declared no competing interest.

### Funding Statement

This research was funded by the South-Eastern Norway Regional Health Authority (Grant
7 Nos. 2016049, 2017025, 2018075 and 2020088 [to AB]), and by Norwegian Research
8 council, #309762 Precision Health Center for optimized cardiac care (ProCardio).

### Author Declarations

The study was approved by the Regional Committees for Medical and Health Research Ethics South East Norway (REC) (2013/601)

