## Supplementary Material for "Severe biventricular cardiomyopathy in both current and former long-term users of anabolic-androgenic steroids"

### SUPPLEMENTAL FIGURES AND TABLES

**Supplemental Figure 1. Compounds used among AAS-users.**

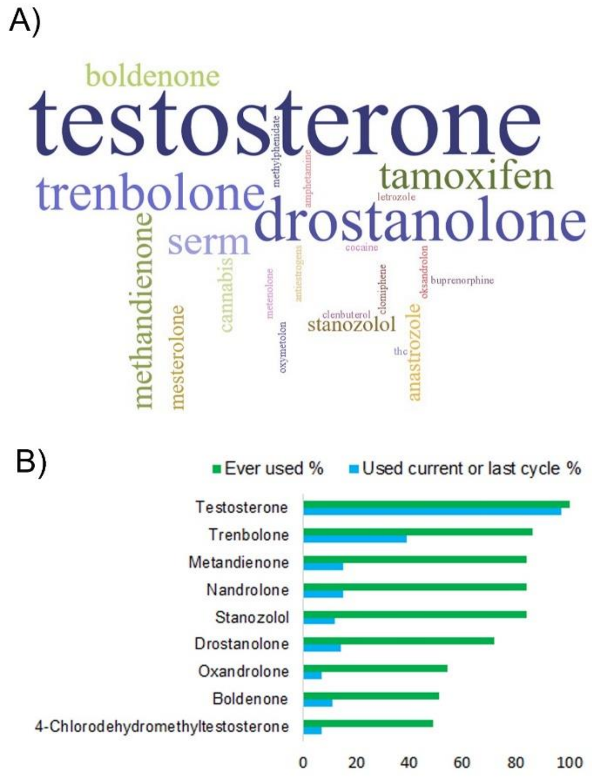


**A)** Analytic findings in urine, including findings of antioestrogens and other substances of abuse, such as opiates, shown as a word cloud. Urine samples were available from 163 participants (9 missing), thereof 95 users. All positive AAS findings originated from the AAS group. Testosterone findings are based upon of samples with testosterone to epitestosterone ratio (T/E) above 15. The top three most frequent findings were testosterone with 47, drostanolone with 29, and trenbolone with 23 positive findings.

**B)** Bar plot of self-reported use of AAS. Green bars are the percentage of participants that have ever used the compound, and the blue bars represent the use of the respective compound in the current or last cycle. Only the mostly used AAS compounds are included, and not growth hormones, insulin, clenbuterol or compounds used to counteract negative side-effects (e.g. antiestrogens), that are instead reported in text. Data are available from 88 AAS-users (13 missing).

**Supplemental Table 1. Adjusted linear regression model comparing key echocardiographic parameters between current and former AAS-users, and WLC.**

| | Current | Former | WLC | F | P | $\eta p^2$ |
|---|---|---|---|---|---|---|
| LVMI ($g/m^2$) | 111±25 | 94±26 | 80±15 | 13,78 | <0.001[a,b] | 0.18 |
| LVEF (%) | 49±7 | 50±7 | 59±5 | 20,36 | <0.001[b,c] | 0.25 |
| LVGLS (%) | -15.0±2.2 | -17.6±2.1 | -18.1±2.2 | 9,43 | <0.001[b] | 0.17 |
| RVGLS (%) | -17. 2±3.5 | -17.4±3.5 | -22.8±2.0 | 22,42 | <0.001[b,c] | 0.35 |

*Data are presented as mean±SD. The regression models were performed with the cardiac measure as dependent variable, group as fixed factor, and age, systolic blood pressure, bench press 1 repetition maximum as continuous covariates. In the models predicting LVGLS and LVEF, LVMI was included as additional covariate. The Bonferroni post hoc test. [a], current users vs. former users; [b], current users vs. WLC; [c], former users vs. WLC.*
*AAS, anabolic-androgenic steroids; LVMI, left ventricular mass index; LVEF, left ventricular ejection fraction; LVGLS, left ventricular global longitudinal strain; RVGLS, right ventricular global longitudinal strain.*

**Supplemental Table 2. Univariate and multivariate linear regression analyses for determinants of increased LVMI (g/m2).**

| | Univariate analysis | | Multivariate analysis | |
|---|---|---|---|---|
| | β (95% CI) | P | β (95% CI) | P |
| Age (years) | 0.04 (-0.11; 0.19) | 0.59 | 0.05 (-0.01; 0.19) | 0.53 |
| History of AAS-use (yes/no) | 0.50 (0.37; 0.63) | <0.001 | 0.33 (0.15; 0.47) | <0.001 |
| Strength training (hours/week) | 0.14 (-0.01; 0.30) | 0.07 | 0.20 (0.05; 0.34) | 0.01 |
| Endurance training (hours/week) | 0.04 (-0.12; 0.20) | 0.60 | | |
| Bench press 1RM (kg) | 0.36 (0.19; 0.49) | <0.001 | 0.16 (-0.00; 0.30) | 0.05 |
| SBP (mmHg) | 0.30 (0.15; 0.44) | <0.001 | 0.17 (0.03; 0.30) | 0.02 |

*LVMI, left ventricular mass index; AAS, anabolic-androgenic steroids; 1RM, one repetition maximum; SBP, systolic blood pressure.*

**Supplemental Table 3. Univariate and multivariate linear regression analyses for determinants of decreased LVEF (%).**

| | Univariate analysis | | Multivariate analysis | |
|---|---|---|---|---|
| | β (95% CI) | P | β (95% CI) | P |
| Age (years) | -0.19 (-0.34; -0.04) | 0.01 | -0.08 (-0.22; 0.07) | 0.28 |
| BSA ($m^2$) | -0.18 (-0.33; -0.03) | 0.02 | 0.00 (-0.16; 0.17) | 0.97 |
| History of AAS-use (yes/no) | -0.62 (-0.74; -0.50) | <0.001 | -0.53 (-0.69; -0.37) | <0.001 |
| Strength training (hours/week) | -0.06 (-0.23; 0.09) | 0.42 | | |
| Endurance training (hours/week) | 0.06 (-0.10; 0.22) | 0.44 | | |
| Bench press 1RM (kg) | -0.31 (-0.47; -0.14) | <0.001 | -0.04 (-0.21; 0.14) | 0.70 |
| SBP (mmHg) | -0.21 (-0.37; -0.06) | 0.006 | -0.01 (-0.16; 0.14) | 0.87 |
| LVMI ($g/m^2$) | -0.41 (-0.59; -0.27) | <0.001 | -0.13 (-0.31; 0.03) | 0.11 |

*LVEF, left ventricular ejection fraction; BSA, body surface area; AAS, anabolic-androgenic steroids; 1RM, one repetition maximum; SBP, systolic blood pressure; LVMI, left ventricular mass index.*

**Supplemental Table 4. Univariate and multivariate linear regression analyses for determinants of decreased LV function by GLS (%).**

| | Univariate analysis | | Multivariate analysis | |
|---|---|---|---|---|
| | β (95% CI) | P | β (95% CI) | P |
| Age (years) | 0.08 (-0.09; 0.26) | 0.35 | 0.03 (-0.15; 0.23) | 0.71 |
| BSA ($m^2$) | -0.04 (-0.21; 0.14) | 0.67 | | |
| History of AAS-use (yes/no) | 0.43 (0.27; 0.58) | <0.001 | 0.24 (0.05; 0.46) | 0.02 |
| Strength training (hours/week) | 0.07 (-0.12; 0.25) | 0.46 | | |
| Endurance training (hours/week) | -0.05 (-0.23; 0.14) | 0.61 | | |
| Bench press 1RM (kg) | 0.34 (0.18; 0.59) | <0.001 | 0.18 (-0.00; 0.41) | 0.06 |
| SBP (mmHg) | 0.26 (0.08; 0.42) | 0.004 | 0.10 (-0.10; 0.31) | 0.31 |
| LVMI ($g/m^2$) | 0.50 (0.35; 0.65) | <0.001 | 0.26 (0.07; 0.47) | 0.008 |

*LV, left ventricle; GLS, global longitudinal strain; BSA, body surface area; AAS, anabolic-androgenic steroids; 1RM, one repetition maximum; SBP, systolic blood pressure; LVMI, left ventricular mass index.*

**Supplemental Table 5. Univariate and multivariate linear regression analyses for determinants of decreased RV function by GLS (%).**

| | Univariate analysis | | Multivariate analysis | |
|---|---|---|---|---|
| | β (95% CI) | P | β (95% CI) | P |
| Age (years) | 0.03 (-0.16; 0.22) | 0.76 | -0.06 (-0.22; 0.10) | 0.47 |
| BSA ($m^2$) | 0.19 (0.00; 0.36) | 0.04 | 0.10 (-0.08; 0.27) | 0.28 |
| History of AAS-use (yes/no) | 0.67 (0.54; 0.82) | <0.001 | 0.65 (-.47; 0.87) | <0.001 |
| Strength training (hours/week) | -0.00 (-0.21; 0.20) | 0.96 | | |
| Endurance training (hours/week) | -0.07 (-0.26; 0.12) | 0.47 | | |
| Bench press 1RM (kg) | 0.35 (0.17; 0.59) | 0.001 | -0.08 (-0.30; 0.13) | 0.44 |
| SBP (mmHg) | 0.20 (0.02; 0.37) | 0.03 | -0.01 (-0.16; 0.148) | 0.95 |
| LVMI ($g/m^2$) | 0.49 (0.32; 0.63) | <0.001 | 0.15 (-0.04; 0.34) | 0.13 |

*RV, right ventricle; GLS, global longitudinal strain; BSA, body surface area; AAS, anabolic-androgenic steroids; 1RM, one repetition maximum; SBP, systolic blood pressure; LVMI, left ventricular mass index.*

**Supplemental Table 6. Univariate linear regression analyses for determinants of decreased LVEF (%), when adding use of psychostimulants as additional covariates.**

| | β (95% CI) | P |
|---|---|---|
| Age (years) | -0.06 (-0.20; 0.09) | 0.45 |
| BSA ($m^2$) | 0.03 (-0.14; 0.19) | 0.74 |
| History of AAS-use (yes/no) | -0.54 (-0.70; -0.34) | <0.001 |
| Bench press 1RM (kg) | -0.02 (-0.19; 0.16) | 0.85 |
| SBP (mmHg) | -0.02 (-0.17; 0.12) | 0.77 |
| LVMI ($g/m^2$) | -0.18 (-0.34; -0.01) | 0.04 |
| Use of amphetamines | -0.01 (-0.16; 0.14) | 0.88 |
| Use of cocaine | 0.02 (-0.14; 0.18) | 0.79 |

*LVEF, left ventricular ejection fraction; BSA, body surface area; AAS, anabolic-androgenic steroids;1RM, one repetition maximum; SBP, systolic blood pressure; LVMI, left ventricular mass index.*

**Supplemental Table 7. Univariate linear regression analyses for determinants of decreased LV function by GLS (%), when adding use of psychostimulants as additional covariates.**

| | β (95% CI) | P |
|---|---|---|
| Age (years) | 0.01 (-0.18; 0.20) | 0.90 |
| History of AAS-use (yes/no) | 0.25 (0.01; 0.50) | 0.04 |
| Bench press 1RM (kg) | 0.15 (-0.05; 0.38) | 0.12 |
| SBP (mmHg) | 0.08 (-0.13; 0.29) | 0.43 |
| LVMI ($g/m^2$) | 0.31 (0.11; 0.58) | 0.01 |
| Use of amphetamines | 0.09 (-0.11; 0.28) | 0.39 |
| Use of cocaine | -0.15 (-0.35; 0.06) | 0.16 |

*LVGLS, left ventricular global longitudinal strain; BSA, body surface area; 1RM, one repetition maximum; SBP, systolic blood pressure; LVMI, left ventricular mass index.*

**Supplemental Table 8. Adjusted linear regression model comparing key echocardiographic parameters between three groups of current AAS-users with varying duration of AAS-use, and WLC.**

| | WLC | Group A (n=8)<br><5 years of use | Group B (n=15)<br>5-10 years of use | Group C (n=32)<br>>10 years of use | P |
|---|---|---|---|---|---|
| LVMI (g/m$^2$) | 82±15 | 107±24* | 104±28** | 113±22*** | <.001 |
| LVEF (%) | 59±5 | 52±6 | 49±7*** | 47±6 *** | <.001 |
| LVGLS (%) | -18.2±2.2 | -15.9±2.0 | -14.8±2.8* | -14.6±2.2 | 0.041 |
| RVGLS (%) | -22.8±2.0 | -17.7±3.2* | -16.5±3.8*** | -16.9±4.0** | 0.017 |

*Data are presented as mean±SD and n(%) when appropriate. The regression models were performed with the cardiac measure as dependent variable, group as fixed factor, and age, systolic blood pressure, bench press 1 repetition maximum as continuous covariates. In the models predicting LVGLS and LVEF, LVMI was included as additional covariate. The Bonferroni post hoc test. * p<.05, ** p<.01, *** p<.001 significantly different from WLC. No differences were found between subgroups of users varying in different length of AAS-use. AAS, anabolic-androgenic steroids; T/E ratio, testosterone to epitestosterone ratio; BSA, body surface area; BMI, body mass index; SBP, systolic blood pressure; DBP, diastolic blood pressure.*
